# boutliers: R package of outlier detection and influence diagnostics for meta-analysis

**DOI:** 10.1101/2025.09.18.25336125

**Authors:** Hisashi Noma, Kazushi Maruo, Masahiko Gosho

## Abstract

Meta-analysis is an established methodology for evidence synthesis. In practice, substantial heterogeneity often arises among studies, and random-effects models are widely employed as standard tools. However, in many cases of data synthesis, some studies exhibit markedly different characteristics from others, beyond the degree expected from statistical error, and may become influential outliers that affect the overall conclusions. Although outlier detection and influence diagnostic methods have been discussed in the context of meta-analysis, there has been a lack of user-friendly statistical packages that quantify the statistical uncertainty of diagnostic measures. We developed the R package `boutliers`, which implements influence diagnostics based on bootstrap methods using simple commands. The package provides three leave-one-out diagnostics: (1) studentized residuals, (2) relative change measures for the variance of the grand mean parameter, and (3) relative change measures for heterogeneity variance. In addition, a model-based approach using a likelihood ratio statistic under a mean-shifted outlier detection model is also available. This article offers a practical tutorial for the `boutliers` package, illustrated with an application to a meta-analysis of chronic low back pain.

## 1. Introduction

Systematic review is an established research method in evidence-based medicine, and meta-analysis serves as its methodological framework for data analysis (Egger et al. 2022; Higgins and Thomas 2019). In the application of meta-analyses, substantial heterogeneities often arise among different studies, e.g., study designs, participant characteristics, regions, sites, treatment administration, interventions, and outcome definitions. To effectively address this heterogeneity, random-effects models are commonly employed as standard tools (Borenstein et al. 2021; Higgins and Thomas 2019). However, in many instances of data synthesis, some studies may exhibit markedly different characteristics from others, surpassing the degree of statistical errors that random-effects models can account for. These studies are formally identified as “outliers” (Harrer et al. 2021; Viechtbauer and Cheung 2010). In general statistical theory, it is well-known that outliers can introduce biases and potentially lead to misleading results, and various statistical theories and analysis tools are developed (Belsley et al. 1980; Cook and Weisberg 1982; Weisberg 2014). Since the evidence created by meta-analysis has been widely used in public health, clinical practices, health technology assessments, and policy making, the outlier problem is a relevant issue that must be addressed to avoid serious impacts in these applications.

A primary approach to tackling the outlier problem involves conventional outlier detection methods developed for ordinary regression analyses (Belsley et al. 1980; Cook and Weisberg 1982; Weisberg 2014). These methods are simple yet powerful tools and are included in standard statistical software (e.g., `PROC REG` in SAS, `lm` function in R). Viechtbauer and Cheung (2010) proposed to adapt these conventional regression diagnostic methods to the meta-analysis framework and their methods have been widely used in meta-analysis practices. Also, these methods were extended to more general methods, e.g., bivariate meta-analysis for diagnostic test accuracy (Negeri and Beyene 2020) and network meta-analysis (Noma et al. 2020). Similar methods have also been discussed for addressing outlier issues in multicenter and multiregional clinical trials (Aoki et al. 2021; Nakamura and Noma 2021).

Although the influence diagnosis methods of Viechtbauer and Cheung (2010) have been widely cited in practice of systematic reviews, a relevant limitation is the lack of certain methods that provide thresholds for influence statistics to quantify the influences of individual studies. Formal thresholds are typically given by quantiles of the normal distribution; however, these represent only rough approximations. Besides, recent studies have proposed using bootstrap methods (Efron and Tibshirani 1994) to quantitatively evaluate statistical variabilities and provide reasonable rationales for how “outlying” candidate studies are within the overall population (Aoki et al. 2021; Nakamura and Noma 2021; Negeri and Beyene 2020; Noma et al. 2020). However, there are currently no user-friendly computational tools readily accessible to non-statisticians for these methods.

In this article, we introduce an R package, `boutliers` available at CRAN (https://doi.org/10.32614/CRAN.package.boutliers), which performs these influence diagnostics based on the bootstrap approach using simple commands. The `boutliers` package can handle three diagnostic statistics using a leave-one-out scheme: (1) studentized residual, (2) relative change measure for the variance of the grand mean parameter, and (3) relative change measure for heterogeneity variance. Additionally, a model-based diagnostic approach using a likelihood-ratio statistic with a mean-shifted outlier detection model is available. These four methods focus on different aspects of characterizing the extremeness of individual studies and will be useful for identifying outlying studies from various perspectives. The remainder of this paper is structured as follows. Section 2 briefly describes these methods, Section 3 provides an overview of the `boutliers` package, and Section 4 provides detailed explanations of individual functions with applications to real-world examples. Concluding remarks are presented in Section 5.

## 2. Methods

### 2.1 Statistical models and inference methods

We consider the standard DerSimonian-Laird-type random-effects model (DerSimonian and Laird 1986) as the basic framework. Let *Y*_*i*_ (*i* = 1, ⋯, *N*) denote the study-specific summary statistics of the treatment effect measures (e.g., mean difference, log risk ratio, log hazard ratio), with corresponding variances 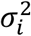. The random-effects model is then expressed as

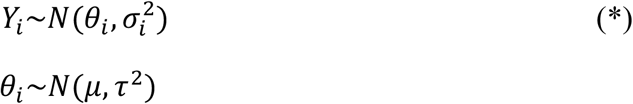

where *μ* is the grand mean parameter and *τ*^2^ is the between-study heterogeneity variance. If all study-specific effect *θ*_*i*_ are equal (*τ*^2^ = 0), this model reduces to the fixed-effect model, which assumes a common effect across the included studies (Higgins and Thomas 2019).

Various methods are available for estimating the model parameters (Veroniki et al. 2019). In this paper, we adopt the inverse-variance method for the fixed-effect model and the restricted maximum likelihood (REML) method for the random-effects model as standard approaches, although other estimation methods may be employed (e.g., the Hartung-Knapp-Sidik-Jonkman (HKSJ) method; IntHout et al. 2014). The log-likelihood function of the random-effects model is given by

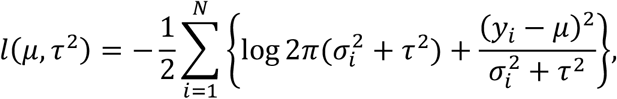

and the log-restricted likelihood function is given by

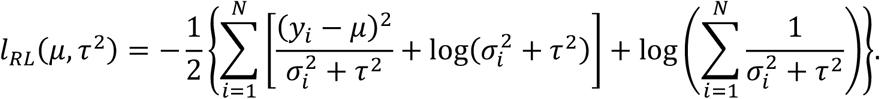

The REML estimator of 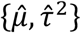 are obtained by maximizing *l*_*RL*_(*μ, τ*^2^). In the following sections, we primarily present influence diagnostic methods based on the random-effects model (*). However, these methods can be straightforwardly adapted to the fixed-effect model, since it corresponds to model (*) with the restriction *τ*^2^ = 0.

### 2.2 Studentized residual

First, we discuss the studentized residual, which is analogous to the DFBETA statistic and has been widely used in conventional regression diagnostics (Belsley et al. 1980; Cook and Weisberg 1982). This is one of the most well-known influence diagnostic measures, defined as the standardized residuals divided by their standard errors, making them comparable across all analysis units. The naive studentized residual for the *a*th study (*a* = 1, ⋯, *N*) is formally defined as

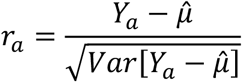

where 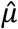 is the REML estimator of *μ*, although it may be replaced with other valid estimators (e.g., the HKSJ method). This influence measure is intuitively useful because it quantifies the divergence of *Y*_*a*_ for individual studies from the overall mean estimate 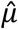 However, it may not be suitable for assessing the influence of individual studies, since it relies on information from the very studies used to estimate the overall mean *μ*; in other words, the evaluation of their influence can involve a degree of “optimism.”

Thus, recent studies have adopted leave-one-out–type measures to circumvent the optimism inherent in these influence measures (Viechtbauer and Cheung 2010). Let 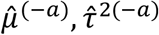 denote the REML estimators from the random-effects model (*) based on the dataset of *N* - 1 studies excluding the *a*th study. Then, the leave-one-out studentized residual is defined as

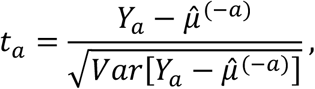

where 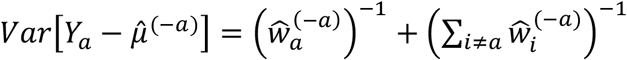 and 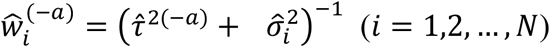, which can be interpreted as the predicted studentized residual of the *a* th study. Note that the estimates 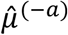 and 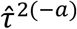 do not depend on the corresponding *a*th study; therefore, this diagnostic measure effectively circumvents the optimism.

To evaluate the influences quantitatively, appropriate reference values are required in practice. Conventionally, the standard normal distribution N(0, 1) has been widely used as an approximate sampling distribution of *r*_*a*_ and *t*_*a*_. Accordingly, a commonly-used criterion is to compare the absolute values of *r*_*a*_ and *t*_*a*_ with 1.96 (or its rounded value of 2.00). If this criterion is met, the corresponding study is typically considered as a potential outlier that exceeds the expected range of statistical variation. However, this rule relies on the large-sample assumptions and may be inadequate in practice. Therefore, we adopt a bootstrap-based approach to quantify sampling variation, leveraging modern computational techniques, as in Aoki et al. (2021) and Nakamura and Noma (2021). The bootstrap algorithm is given as follows.

#### Algorithm 1

*(Bootstrap for estimating the sampling distribution of t_a_)*.

1. For the random effects model (*), compute the REML estimates of 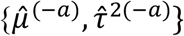 based on the dataset of *N*- 1 studies excluding the *a*th study.
2. Perform parametric bootstrap resampling 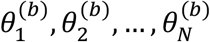 from the estimated distribution 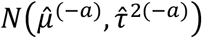 for *b* = 1,2,…,*B*. Also, generate resampled data 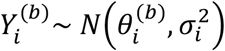 for *i* = 1,2, …, *N*.
3. Compute the leave-one-out studentized residuals 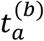 for the *b*th bootstrap sample 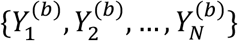.
4. The bootstrap estimate of the sampling distribution of *t*_*a*_ is obtained from the empirical distribution of 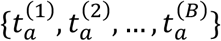.

We can then provide reasonable thresholds from the bootstrap-based estimated sampling distribution. For example, the 2.5th and 97.5th percentiles of the empirical distribution can be adopted as thresholds, and studies exceeding these values may be considered influential outliers beyond the expected range of statistical variation. The same method can also be applied to the fixed-effect model by setting *τ*^2^ = 0. In the `boutliers` package, these calculations can be performed with the functions `STR` using simple commands.

### 2.3 Relative change of the variance of the overall estimator of μ

Second, we consider an influence measure defined as the relative change in the variance estimate of the overall estimator of *μ* under the leave-one-out scheme. This measure was originally proposed by Viechtbauer and Cheung (2010) and is defined as

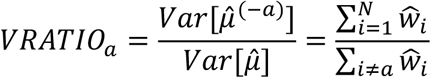

This measure assesses the relative change in variance between the leave-one-out dataset and the dataset containing all studies. *VRATIO*_*a*_ reflects the impact of the *a*th study on the precision of the overall treatment effect estimator. The values of *VRATIO*_*a*_ lie in (0, ∞). When *VRATIO*_*a*_ is close to 1, the variance estimate remains nearly unchanged even if the *a*th study is excluded, indicating that the study is not influential for the precision of the variance estimator.

If *VRATIO*_*a*_ is much smaller than 1, exclusion of the *a*th study substantially decreases the uncertainty of the overall mean estimator, despite the reduced sample size. In such cases, the study may be considered influential in the sense that its inclusion inflates the uncertainty—typically because an outlying study deviates from the overall mean and increases variance. These studies can therefore be candidates for influential outliers.

Conversely, when *VRATIO*_*a*_ is larger than 1, exclusion of the *a*th study increases the uncertainty of the overall estimator. This is natural, as the sample size is reduced, and the increase in variance is usually not large. If *VRATIO*_*a*_ takes a very large value, the corresponding study may possess unique characteristics, but it would not typically be regarded as an influential outlier in the standard sense.

To quantify the degree of outlyingness, the bootstrap algorithm described in Section 2.2 can be applied to calculate the bootstrap distribution of *VRATIO*_*a*_ by substituting *t*_*a*_ with *VRATIO*_*a*_ in Algorithm 1 (Aoki et al. 2021; Nakamura and Noma 2021). For example, the lower 5th percentile of the bootstrap distribution can be used as a critical value.

For the fixed-effect model, however, variance ratio statistics usually reflect the amount of statistical information contributed by each study, which is simply proportional to the inverse of its within-study variance. Therefore, this measure is not adequate for assessing influence under the fixed-effect model and should be used only with the random-effects model. In the `boutliers` package, this influence measure can be calculated using the `VRATIO` function.

### 2.4 Relative change of the heterogeneity variance τ^2^

Another influence measure similar to *VRATIO*_*i*_ is the relative change in the heterogeneity variance estimate 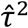. Viechtbauer and Cheung (Viechtbauer and Cheung 2010) proposed using the ratio of the *τ*^2^ estimates obtained from the leave-one-out dataset and from the full dataset.

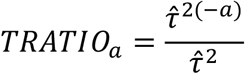

where 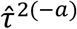 is the REML estimate based on the leave-one-out dataset. The values of *TRATIO*_*a*_ lie in (0, ∞), and its interpretation is similar to that of *VRATIO*_*a*_. If *TRATIO*_*a*_ is approximately 1 or larger, the *a*th study has little influence. Conversely, if *TRATIO*_*a*_ is much smaller than 1, exclusion of the *a*th study decreases between-study heterogeneity and the study may be interpreted as a potential outlier among the *N* synthesized studies.

To quantify the degree of influence, the bootstrap approach (Algorithm 1) can also be applied to estimate the sampling distribution of *TRATIO*_*a*_ (Aoki et al. 2021; Nakamura and Noma 2021). An appropriate criterion is to use the lower 5th percentile of the bootstrap distribution as the critical value.

Note that this measure cannot be defined for the fixed-effect model and can only be applied under the random-effects model. As discussed by Viechtbauer and Cheung (2010), this measure serves as an effective influence diagnostic when substantial heterogeneity is present.

### 2.5 Model-based approach using a mean-shifted model

Another commonly used approach to influence diagnostics is a model-based method that compares the original model with an alternative model that explicitly accounts for potential outliers. Aoki et al. (2021) and Nakamura and Noma (2021) adopted a mean-shifted model, which assumes that the treatment effect of one study among the *N* studies differs substantially from the others.

For the DerSimonian–Laird-type random-effects model (*), we consider that the random-effect distribution for the corresponding *a*th study is shifted as

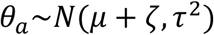

while that of the remaining *N*-1 studies follows the same distribution as in (*), i.e.,

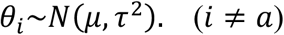

We then consider the following hypothesis test:

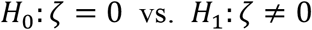

If the null hypothesis is rejected, the treatment effect of the *a*th study is significantly different from the overall mean and the study may be considered potentially influential. Note, however, that the alternative hypothesis represents a nonzero mean difference and does not necessarily imply that the corresponding study is an outlier with an extreme effect among the synthesized studies. Therefore, the purpose of this model-based approach should be interpreted not as “outlier detection,” but rather as “detection of an influential study.”

In addition, the statistical power of this test is usually limited; therefore, studies that are significantly detected may indeed have large impacts and might be outliers, but interpretation should be made carefully in the context of the specific research field, ideally in consultation with subject-matter experts.

For this testing problem, a straightforward approach is the likelihood-ratio test (Aoki et al. 2021; Nakamura and Noma 2021). The log-likelihood function under *H*_O_ corresponds to that of the random-effects model (*):

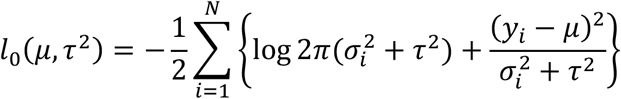

The log-likelihood function under *H* is given by

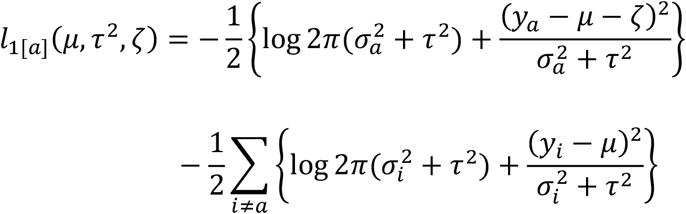

The likelihood-ratio statistic is then written as

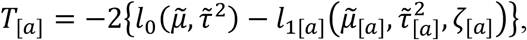

where 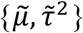 is the maximum likelihood (ML) estimates under the null model, and 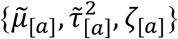 are the ML estimates under the mean-shifted model for the *a*th study.

According to conventional large-sample theory, the likelihood-ratio statistic *T*_[a]_ approximately follows a *χ*^2^ distribution with 1 degree of freedom under the null hypothesis. However, as discussed in the previous sections, this large-sample approximation may be violated in realistic settings, and the chi-square approximation can be unreliable (Noma 2011; Noma et al. 2018). For this model-based approach, a bootstrap-based procedure is also applicable.

#### Algorithm 2

*(Bootstrap test for the model-based likelihood-ratio statistic)*.

1. Compute the ML estimates 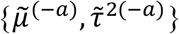 based on the dataset of *N* - 1 studies excluding the *a*th study.
2. Perform parametric bootstrap resampling *B* times (*b* = 1,2,…,*B*). Specifically, generate samples 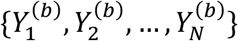 from the estimated null model (*), where the parameters are set to 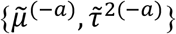.
3. For each bootstrap sample 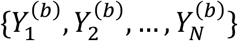, compute the ML estimates 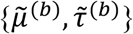 under the null model and 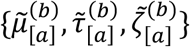 under the mean-shifted model. Then calculate the likelihood-ratio statistic:

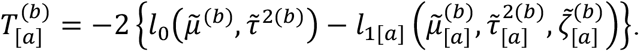 Repeat this step for all *B* bootstrap samples.
4. Construct the bootstrap distribution of *T*_[*a*]_ from the empirical distribution of 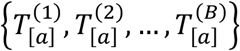 The bootstrap *p*-value is then obtained from this bootstrap distribution.

In summary, the likelihood ratio test serves as a useful diagnostic tool for identifying influential studies. Combined with bootstrap calibration, it enables more reliable inference in realistic meta-analytic settings.

## 3. R package *boutliers*

`boutliers` is an R package available on CRAN. It implements influence diagnostics for meta-analysis models, as introduced in the previous section. Table 1 summarizes the available functions, each corresponding to one of the methods previously described:

- `STR`: influential analysis based on studentized residuals
- `VRATIO`: influential analysis based on relative changes in variance estimates
- `LRT`: likelihood-ratio test based on the mean-shifted model

**Table 1.**
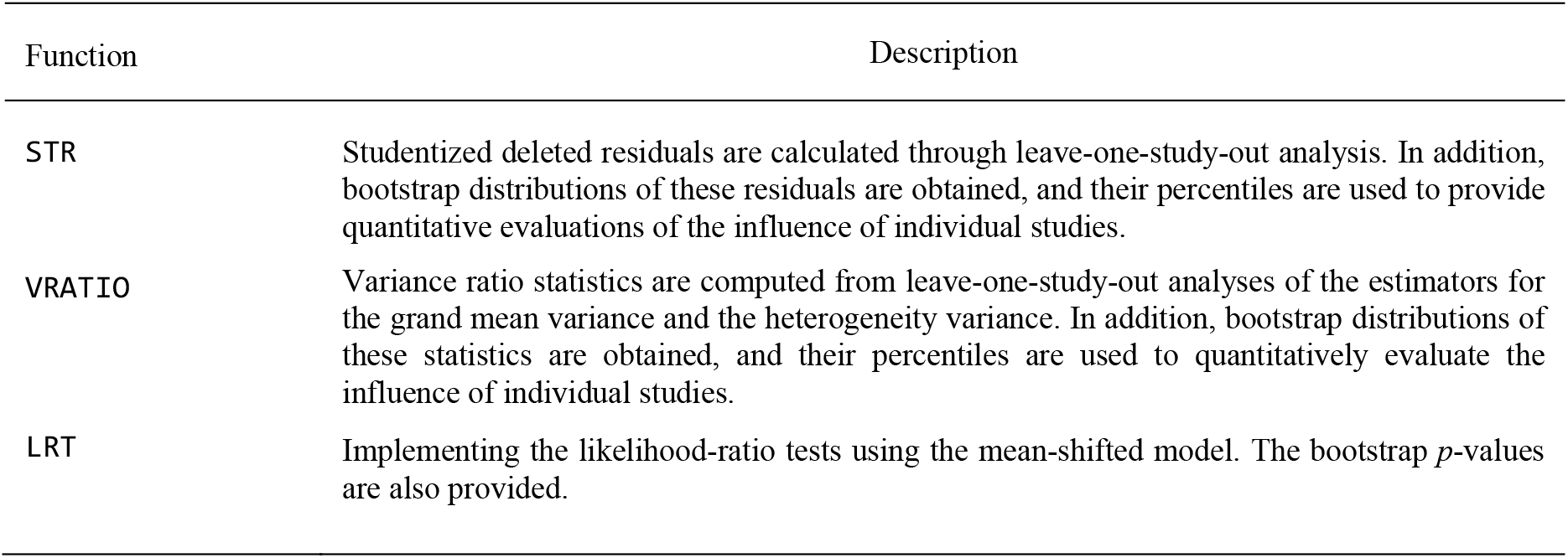
Description of functions contained in boutliers package.

| Function | Description |
| --- | --- |
| STR | Studentized deleted residuals are calculated through leave-one-study-out analysis. In addition, bootstrap distributions of these residuals are obtained, and their percentiles are used to provide quantitative evaluations of the influence of individual studies. |
| VRATIO | Variance ratio statistics are computed from leave-one-study-out analyses of the estimators for the grand mean variance and the heterogeneity variance. In addition, bootstrap distributions of these statistics are obtained, and their percentiles are used to quantitatively evaluate the influence of individual studies. |
| LRT | Implementing the likelihood-ratio tests using the mean-shifted model. The bootstrap $p$-values are also provided. |

All functions can be executed with simple commands, and their syntax is designed to be easily accessible even to non-statisticians. Detailed explanations and illustrations are provided in Section 4 through case studies. The package can be installed and loaded as follows:

~~~
> install.packages(“boutliers”)
> library(“boutliers”)
~~~

The implemented methods are also applicable to detecting influential centers or regions in multi-center or multi-regional clinical trials (Aoki et al. 2021; Nakamura and Noma 2021). To facilitate such applications, the package includes example datasets representing these types of data. All R example programs used in this article can be accessed at https://github.com/nomahi/boutliers/blob/master/Examplecode.r

## 4. Functionality and features with illustrative examples

### 4.1 Example dataset: Meta-analysis of chronic low back pain

As an illustrative example, we consider the meta-analysis by Rubinstein et al. (Rubinstein et al. 2019), who evaluated the benefits and harms of spinal manipulative therapy (SMT) for patients with chronic low back pain. This dataset, provided as `SMT` in the `boutliers` package, includes 23 randomized controlled trials. Figure 1 presents the forest plot of their synthesis, where the outcome was pain intensity at one month (0–100 scale, with higher values indicating worse pain). The comparison was SMT (N = 1,629) versus guideline-recommended therapies (N = 1,526), with the mean difference (MD) as the effect measure.

**Figure 1.**
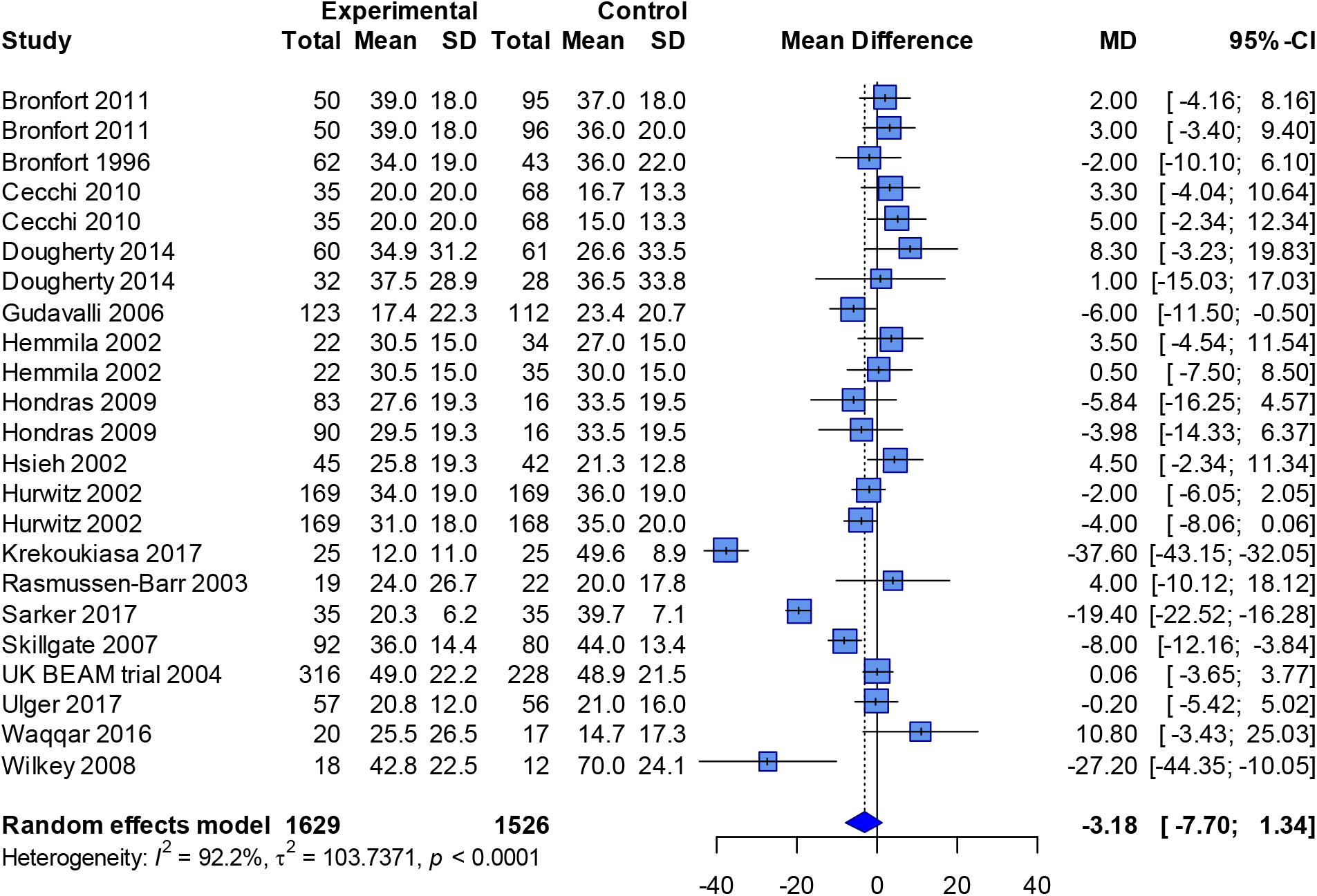
Forest plot for the meta-analysis of chronic low back pain (Rubinstein et al. 2019).

Using the DerSimonian–Laird random-effects model (DerSimonian and Laird 1986), the pooled MD was −3.18 (95% CI: −7.70 to 1.34), with substantial heterogeneity (*I*^2^ = 92%, *τ*^2^ = 103.74, *P* < 0.01, Cochran’s Q-test). Although most trial-specific MDs were close to the pooled mean, a few studies reported much larger effects. These results suggest the presence of influential outliers that may markedly affect the overall estimate. To ensure valid inference about the effectiveness of the intervention, their influence must be quantified accurately. The diagnostic tools implemented in `boutliers` are designed precisely for this purpose.

### 4.2 Studentized residuals

Influence analysis using studentized residuals can be performed with the `STR` function. The arguments of the `STR` function and its output are summarized in Table 2. The function operates with simple syntax: prepare a data frame containing the variables required for the DerSimonian–Laird random-effects model (a vector of the estimated effect size and its estimated variance) and specify this data frame as `data`. Then, by specifying the variable names corresponding to the estimated effect size and its estimated variance as `y` and `v`, respectively (these can, for example, be easily calculated using the `escalc` function from the `metafor` package (Viechtbauer 2010), the function will automatically perform a leave-one-study-out analysis for all studies. The number of bootstrap resampling iterations can be set using `B` (default: 2000), and the percentiles of the bootstrap distribution to be output can be specified with `alpha` (by default, the 2.5th and 97.5th percentiles are used). Additionally, to ensure the results from the bootstrap method are reproducible, a random seed can be specified with `seed`.

**Table 2.** Description of the functionalities of the STR function.

|  | Description |
| --- | --- |
| **Arguments** |  |
| **y** | A vector of the outcome measure estimates (e.g., MD, SMD, log OR, log RR, RD, log HR). |
| **v** | A vector of the variance estimate of **y**. |
| **method** | A logical value specifying the estimation method (default: REML). The same options as those available for the **method** argument of the **rma** function in the **metafor** package can be used (e.g., FE for the fixed-effect model, SJ for the Sidik–Jonkman method, and PM for the Paule–Mandel method). |
| **data** | An optional data frame containing the variables **y** and **v**. |
| **B** | The number of bootstrap resampling (default: 2000) |
| **alpha** | The bootstrap percentiles to be outputted; $0.5(1-\alpha)$th and $(1-0.5(1-\alpha))$th percentiles. Default is 0.95; 2.5th and 97.5th percentiles are calculated. |
| **seed** | A numeric value that determines the random seed for reproducibility (default: 123456). |
| **Value** |  |
| **id** | ID of the study. |
| **psi** | The studentized residuals by leave-one-out analysis (studentized deleted residuals). |
| **Q1** | $0.5(1-\alpha)$th percentile for the bootstrap distribution of the studentized residual (default: 2.5th percentile). |
| **Q2** | $1-0.5(1-\alpha)$th percentile for the bootstrap distribution of the studentized residual (default: 97.5th percentile). |

Within the `STR` function, the estimation method for the variance component parameter can be also specified using the `method` argument. The default is the REML method. For the pooling analyses when calculating studentized residuals, the `STR` function calls the `rma` function from the `metafor` package (Viechtbauer 2010). The estimation method for the variance component can be flexibly specified according to the `method` argument of the `rma` function. For example, by setting it to `FE`, a fixed effect model can be used. Also, the recent Cochrane Handbook (Higgins and Thomas 2019) recommends the HKSJ method (IntHout et al. 2014), and the Sidik-Jonkman method (Sidik and Jonkman 2005) can be used by specifying `SJ`. Similarly, the Paule-Mandel method (Paule and Mandel 1982) can be used by specifying `PM`.

The output consists of a list of leave-one-study-out studentized residuals for each study, ordered by their magnitude, and the corresponding percentiles of the bootstrap distribution. The `id` represents the study ID, and `psi` gives the value of the studentized residual. `Q1` and `Q2` are the 0.5×(1−`alpha`)th and 1−0.5×(1−`alpha`)th percentiles of the bootstrap distribution, respectively; if the value of `psi` falls outside these percentiles, the corresponding study may be considered an influential outlier, accounting for statistical error.

#### Example

Analysis using the `STR` function can be performed with the following simple command; the variance component is estimated using the standard REML method.

~~~
> STR(yi, vi, data = edat1, B = 2000)
    id             psi           Q1          Q2
    16     -5.10452658    -1.983772    1.912068
    23     -1.85339334    -2.017066    1.907075
    18     -1.69817290    -1.898156    1.869712
    22      1.14244992    -1.945369    2.060744
     6      0.99582436    -1.925249    1.905155
     5      0.76418437    -1.861517    1.870008
    13      0.72202878    -1.972032    1.949469
     9      0.61341890    -1.929967    1.981855
     4      0.60182407    -1.938742    1.923383
     2      0.58188314    -1.946533    1.924879
    17      0.58020717    -1.775979    1.869691
     1      0.48784327    -1.905836    1.971817
    19     -0.46496792    -1.870574    1.887666
    10      0.33570141    -1.914289    1.842388
     7      0.32153949    -1.957926    1.732499
    20      0.31204256    -1.927149    1.838943
    21      0.28243298    -1.863396    1.947016
     8     -0.26731755    -1.898940    1.882677
    11     -0.23247766    -1.919655    1.907316
    14      0.11254530    -1.857791    2.057090
     3      0.10678827    -2.028355    1.935564
    15     -0.07930223    -1.827371    1.882818
    12     -0.07029222    -1.941368    1.831976
~~~

The output is ranked according to the magnitude of `psi`. `Q1` and `Q2` correspond to the 2.5th and 97.5th percentiles of the bootstrap distribution. Conventionally, a value exceeding 1.96 or 2.00 was used to determine whether an individual study is an influential outlier, but using this method, more precise thresholds based on bootstrap can be provided. According to these results, the 16th study has been identified as an influential outlier. This study shows an extremely large effect size within the overall distribution, and influences the overall synthesis result. Similarly, by adding an argument such as `method=`“`FE`” or `method=`“`SJ`”, the model or method used for estimation can be changed; these methods should be specified according to the primary statistical analysis method.

### 4.3 Variance ratio statistics

Influence analysis using variance ratio statistics (variance of estimators and heterogeneity variance) can be performed with the `VRATIO` function. The arguments and outputs used in the `VRATIO` function are summarized in Table 3. The `VRATIO` function is also a function that operates with simple syntax. For this function as well, a data frame that contains the variables used in the DerSimonian-Laird random effects model (a vector with the estimated values of the effect size as the outcome and their estimated variances) should be prepared. Then, by specifying the variable names corresponding to the estimated effect size and its variance in the dataset as `y` and `v`, the function performs leave-one-study-out analyses for all studies. The number of bootstrap resampling iterations can be specified with `B` (default: 2000), and the `alpha` argument specifies the percentile of the bootstrap distribution to output (default is the 5th percentile). In addition, to allow for exact replication of bootstrap results, the random seed can be specified with the `seed` argument. In the `VRATIO` function, the `method` argument also allows the user to specify the method for estimating the variance component parameter in the same manner as the `STR` function.

**Table 3.** Description of the functionalities of the VRATIO function.

|  | Description |
| --- | --- |
| **Arguments** |  |
| **y** | A vector of the outcome measure estimates (e.g., MD, SMD, log OR, log RR, RD, log HR). |
| **v** | A vector of the variance estimate of **y**. |
| **method** | A logical value specifying the estimation method (default: REML). The same options as those available for the method argument of the `rma` function in the `metafor` package can be used (e.g., FE for the fixed-effect model, SJ for the Sidik–Jonkman method, and PM for the Paule–Mandel method). |
| **data** | An optional data frame containing the variables **y** and **v**. |
| **B** | The number of bootstrap resampling (default: 2000) |
| **alpha** | The bootstrap percentile to be outputted (default: 0.05). |
| **seed** | A numeric value that determines the random seed for reproducibility (default: 123456). |
| **Value** |  |
| **id** | ID of the study. |
| **VR** | The VRATIO statistic (relative change of the variance of the overall estimator) by leave-one-out analysis. |
| **Q1** | **alpha**th percentile for the bootstrap distribution of the VRATIO statistic. |
| **TR** | The T2RATIO statistic (relative change of the heterogeneity variance) by leave-one-out analysis. |
| **Q2** | **alpha**th percentile for the bootstrap distribution of the T2RATIO statistic. |

The output provides a list of leave-one-study-out VRATIO and TRATIO for each study, sorted by the magnitude of the variance ratio statistics, along with the percentiles of their bootstrap distributions. The `id` represents the study ID, `VR` represents the VRATIO value, and `TR` represents the TRATIO value. `Q1` and `Q2` correspond to the `alpha`th percentiles of each bootstrap distribution; if the values of `VR` or `TR` fall outside these percentiles, that study can be considered an influential outlier, accounting for statistical error.

#### Example

Analysis using the `VRATIO` function can be executed with a simple command as shown below; the variance component is estimated using the standard REML method.

~~~
> VRATIO(yi,vi,data=edat1,B=2000)
$VRATIO
 id        VR         Q1
 16 0.4778609  0.8862575
 18 0.9616509  0.8924325
 23 0.9649587  0.9395354
 22 1.0233935  0.9240738
  6 1.0392249  0.9198538
 17 1.0600678  0.9389967
  7 1.0633385  0.9405035
  5 1.0664842  0.9058223
 13 1.0708353  0.8922777
  9 1.0755569  0.9085626
  4 1.0780997  0.9033286
  2 1.0816855  0.8892902
 11 1.0854283  0.9243461
 12 1.0877251  0.9021208
  1 1.0878236  0.9043062
 10 1.0896150  0.9105157
  3 1.0949439  0.8935893
 19 1.0951284  0.8964138
 21 1.0992660  0.9024590
  8 1.1000342  0.9066720
 20 1.1014066  0.8955201
 14 1.1060562  0.8957077
 15 1.1067515  0.9024092
$TAU2RATIO
 id        TR        Q2
 16 0.3774660 0.7671174
 18 0.8994492 0.8143563
 23 0.9273419 0.8990506
 22 0.9869376 0.8787000
 6  0.9991795 0.8641722
 5  1.0212304 0.8422518
 13 1.0251690 0.8278997
 17 1.0273825 0.8936548
 9  1.0325444 0.8450526
 4  1.0340808 0.8440986
 7  1.0346205 0.8991170
 2  1.0364291 0.8244430
 1  1.0428474 0.8367700
 10 1.0480181 0.8509361
 11 1.0480773 0.8675458
 19 1.0481061 0.8290497
 12 1.0504878 0.8473187
 21 1.0540856 0.8315141
 3  1.0541042 0.8333208
 20 1.0545496 0.8227138
 8  1.0553381 0.8404581
 14 1.0600855 0.8273693
 15 1.0608702 0.8341120
~~~

The outputs are ranked according to the sizes of `VR` and `TR`. `Q1` and `Q2` correspond to the 5th percentiles of the bootstrap distributions for `VR` and `TR`, respectively. According to these results, as with the studentized residuals, the 16th study is detected as an influential outlier. This can be interpreted as an influential outlier in terms of changes in the estimated variances, after accounting for statistical error.

### 4.4 Likelihood-ratio test under a mean-shifted model

Influence analysis based on the likelihood-ratio test using the mean-shifted model can be conducted with the `LRT` function. The arguments and outputs used in the `LRT` function are summarized in Table 4. The `LRT` function is also a function that operates with simple syntax. For this function as well, a data frame containing the variables used in the DerSimonian-Laird random effects model (a vector consisting of estimated values of the outcome effect size and their estimated variances) should be prepared and specify that dataframe as the `data` argument. Then, by specifying the variable names corresponding to the estimated effect sizes and their variances in the dataset as `y` and `v`, the function will analyze all studies using the likelihood-ratio test based on the mean-shifted model. The number of bootstrap resampling iterations is specified with `B` (default: 2000), and the significance level is specified with `alpha` (default: 0.05). In addition, to allow for exact replication of the bootstrap resampling, a random seed is specified by the `seed` argument. Because the likelihood-ratio test is based on likelihood-based inference, model parameters are estimated using the maximum likelihood estimators. Accordingly, the `LRT` function includes an option to specify whether to use a fixed effects model or a random effects model via the `model` argument (`FE`: fixed-effect model, `RE`: random-effects model); the default is the random-effects model. As output, the function provides the value of the likelihood-ratio statistic for each study, sorted by the magnitude of the likelihood-ratio statistic, the 1−`alpha`th percentile of its bootstrap distribution, and a list of bootstrap *p*-values. `id` represents the study ID, `LR` is the value of the likelihood-ratio statistic, `Q` is the 1−`alpha` percentile of the bootstrap distribution, and `P` is the *p*-value. If significant results are obtained, the corresponding studies can be considered influential outliers, accounting for statistical error.

**Table 4.**
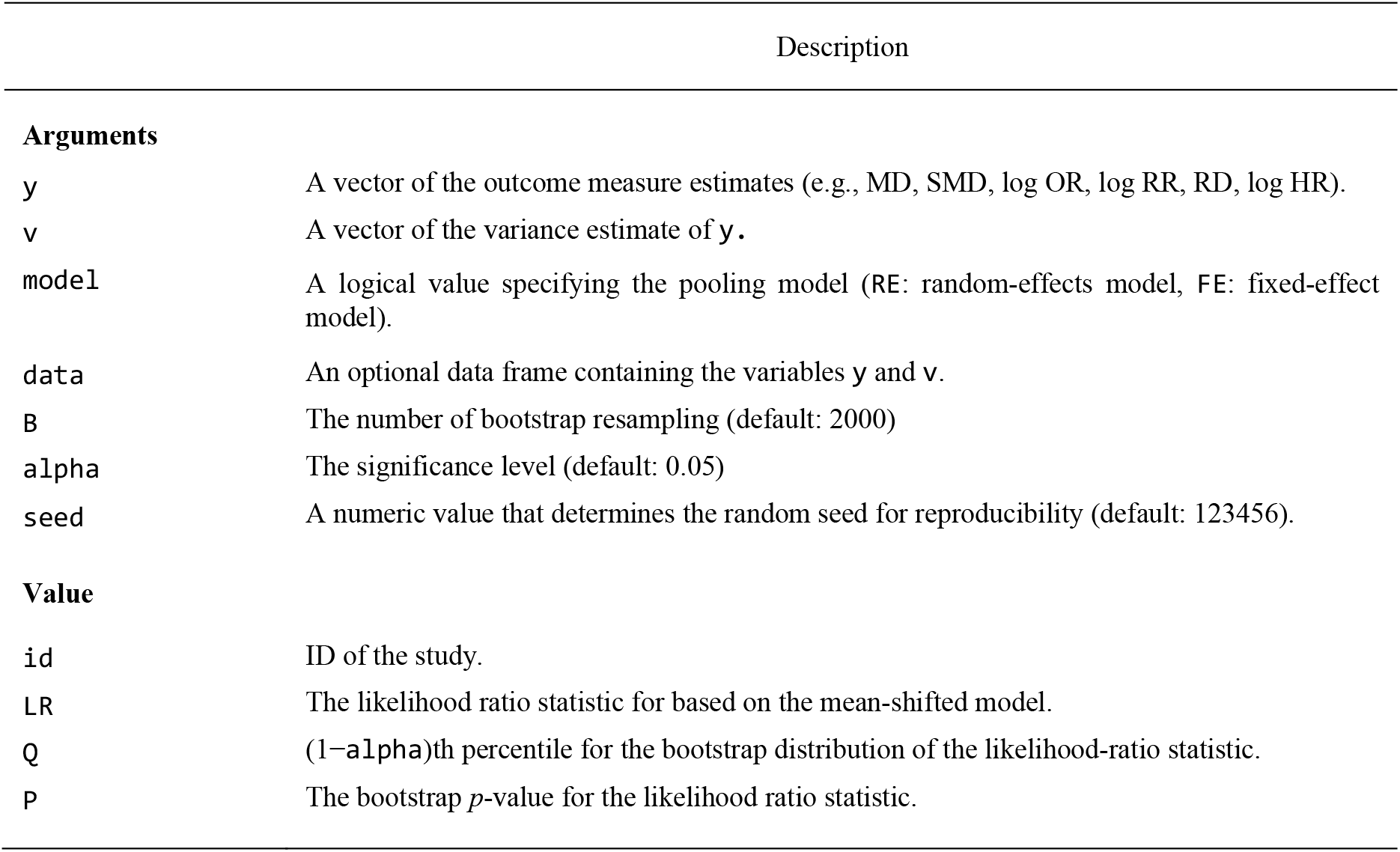
Description of the functionalities of the LRT function.

#### Example

Analysis using the `LRT` function can be executed with a simple command as shown below.

~~~
> LRT(yi,vi,data=edat1,B=2000)
  id           LR         Q           P
  16 17.711008445  4.452721  0.00000000
  23  3.488316594  4.124637  0.06896552
  18  2.966951627  4.512144  0.11344328
  22  1.363317469  4.207158  0.27686157
   6  1.049898689  4.309350  0.34232884
   5  0.634312298  4.258880  0.44777611
  13  0.568438949  4.644110  0.48875562
   4  0.396097527  4.301775  0.56071964
  17  0.357608558  3.779455  0.56121939
   9  0.410062717  4.262856  0.56471764
   2  0.372060122  4.606639  0.59320340
   1  0.262781345  4.293199  0.64517741
  19  0.237411147  4.402671  0.66666667
   7  0.109729841  3.767097  0.74162919
  10  0.124187271  4.151926  0.74312844
  20  0.109382004  4.373924  0.76211894
  21  0.089273691  4.284432  0.77961019
   8  0.077983155  4.317004  0.80209895
  11  0.057697644  4.019434  0.80609695
   3  0.012850919  4.575161  0.90004998
  14  0.014537912  4.363172  0.90154923
  15  0.006690266  4.342537  0.94252874
  12  0.005126409  4.399962  0.94702649
~~~

The output is ranked according to the size of the `LR`. `Q` corresponds to the 95th percentile of the bootstrap distribution and represents the rejection threshold value. According to these results, as in the previous sections, the 16th study is significant and has been detected as an influential outlier. In terms of influence on the estimator of the grand mean, this study can be considered a highly influential outlier. By adding the argument `model=`“`FE`” to this function, it is possible to use a fixed-effects model; the model should be specified to be consistent with the primary statistical analysis method.

Through the above analyses, the 16th study was consistently detected as an influential outlier. Conducting a sensitivity analysis excluding this study, the estimated grand mean and confidence interval based on the DerSimonian-Laird model were −1.61 (95% CI: −4.74, 1.51) (*I*^2^ = 80%, *τ*^2^ = 39.16, *P* < 0.01 by Cochrane’s Q-test). Heterogeneity becomes visibly smaller, and the effect size is also considerably more restrained. This insight is likely to have a significant impact on the interpretation and conclusions of this systematic review.

## 5. Concluding remarks

Meta-analysis is an established method in modern evidence-based medicine and has been widely used in relevant decision-making processes, including the development of clinical guidelines, health technology assessments, and healthcare policy (Egger et al. 2022; Higgins and Thomas 2019). However, it is practically impossible for all studies included in a systematic review to have been conducted under identical protocols, methods, and settings. Accordingly, at least some degree of heterogeneity among studies inevitably influences the interpretation and conclusions of the review. In particular, when studies with markedly divergent profiles are included, which could affect the overall conclusions, interpretation with caution is especially warranted.

Conventional influence diagnostics based on regression techniques, such as those proposed by Viechtbauer and Cheung (2010), are valuable and widely applied in practice. Nevertheless, quantitative tools for identifying influential outliers remain limited. By incorporating bootstrap techniques, it becomes possible to more precisely evaluate statistical errors. The `boutliers` package introduced in this paper provides an efficient and user-friendly tool for conducting such analyses with simple commands. We hope that this package will support researchers and practitioners in improving meta-analysis practice and contribute to the production of evidence that is more relevant for clinical practice and public health.

In the future, integration of influence diagnostics into routine systematic review workflows could enhance transparency and robustness in evidence synthesis. Continued methodological research will also be essential to refine these tools and expand their applicability across diverse clinical contexts.

## Data Availability

All data produced are available online at the CRAN page of boutliers package.

https://doi.org/10.32614/CRAN.package.boutliers

## Disclosure Statement

The author declares no conflicts of interest regarding this article.

## Funding

This study was supported by a Grant-in-Aid for Scientific Research from the Japan Society for the Promotion of Science (grant numbers: JP22H03554, JP23K11931, and JP24K21306).

## Notes

### Competing Interest Statement

The authors have declared no competing interest.

